# Integrated Explainable Ensemble Machine Learning Prediction of Injury Severity in Agricultural Accidents

**DOI:** 10.1101/2025.02.05.25321769

**Authors:** Omer Mermer, Eddie Zhang, Ibrahim Demir

## Abstract

Agricultural injuries remain a significant occupational hazard, causing substantial human and economic losses worldwide. This study investigates the prediction of agricultural injury severity using both linear and ensemble machine learning (ML) models and applies explainable AI (XAI) techniques to understand the contribution of input features. Data from AgInjuryNews (2015–2024) was preprocessed to extract relevant attributes such as location, time, age, and safety measures. The dataset comprised 2,421 incidents categorized as fatal or non-fatal. Various ML models, including Naïve Bayes (NB), Decision Tree (DT), Support Vector Machine (SVM), Random Forest (RF), and Gradient Boosting (GB), were trained and evaluated using standard performance metrics. Ensemble models demonstrated superior accuracy and recall compared to linear models, with XGBoost achieving a recall of 100% for fatal injuries. However, all models faced challenges in predicting non-fatal injuries due to class imbalance. SHAP analysis provided insights into feature importance, with age, gender, location, and time emerging as the most influential predictors across models. This research highlights the effectiveness of ensemble ML models in injury prediction while emphasizing the need for balanced datasets and XAI techniques for actionable insights. The findings have practical implications for enhancing agricultural safety and guiding policy interventions.

**Highlights:**

- This study analyzed 2,421 agricultural injury incidents from AgInjuryNews (2015– 2024) and utilized machine learning models to predict injury severity, focusing on both fatal and non-fatal outcomes.
- Ensemble models, such as XGBoost and Random Forest, outperformed linear models in accuracy and recall, especially in predicting fatal injuries, although challenges in non-fatal predictions due to class imbalance were observed.
- Key predictors identified through SHAP analysis included age, gender, location, and time, providing interpretable insights into the factors influencing injury severity.
- The integration of explainable AI (XAI) enhanced the transparency of machine learning predictions, enabling stakeholders to prioritize targeted safety interventions effectively.
- This research highlights the potential of combining ensemble ML models with XAI techniques to improve agricultural safety practices and provides a foundation for addressing data challenges in future studies.

**Graphical Abstract:** 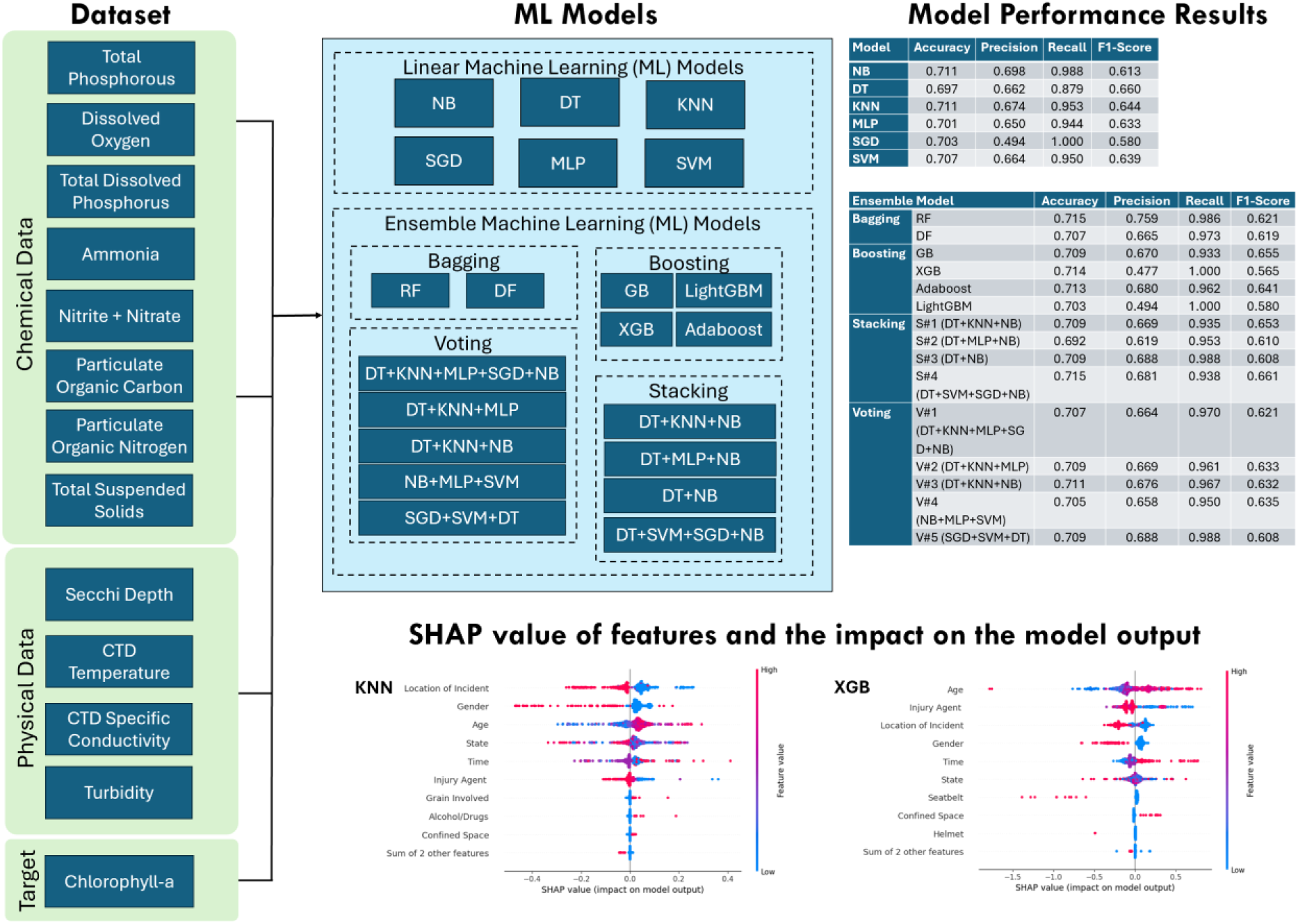

## 1. Introduction

Occupational incidents can significantly impact workers’ lives, both on and off the job, and impose a substantial economic burden on employers, employees, insurance companies, medical care systems, and society as a whole (Suárez Sánchez et al., 2011). The International Labor Organization estimates that nearly 337 million occupational incidents are reported globally each year (Organization, 2008). These incidents result from multiple factors that contribute to their occurrence (Sarkar et al., 2018). Given the vast human and financial losses associated with occupational injuries, researchers have continuously sought to better understand the factors influencing incident occurrence and severity, as well as to improve the accuracy of predicting future injuries (Lord and Mannering, 2010).

Agriculture is a high-risk occupational environment. The agricultural sector consistently ranks among the most hazardous for workers in the U.S., with 23.5 injuries per 100,000 workers reported in 2022 (U.S. Bureau of Labor Statistics, 2023). To reduce this injury rate, stakeholders require detailed information about the nature of these injuries, enabling the development and evaluation of targeted interventions and policies. However, formal reporting requirements in this industry remain relatively minimal, resulting in limited and incomplete national datasets.

Agricultural injury surveillance data plays a crucial role in understanding the circumstances surrounding incidents, aiding prevention efforts. Previous agricultural safety research has focused on collecting, coding, and analyzing injury data from various sources, such as surveys, news reports, workers’ compensation data, and electronic health records. Despite these efforts, agricultural safety and health researchers continue to face challenges in documenting nonfatal injuries, accessing public data systems for surveillance, and assessing the cost-effectiveness of potential data sources or methods (Murphy et al., 2019). Agricultural injury surveillance databases often include basic fields for reporting incident details, such as victim demographics and injury variables. However, manually extracting variables from narrative descriptions is both costly and time-consuming. Measure (2014) estimates that the annual effort required to initially code incidents reported in the Survey of Occupational Injury and Illness (SOII) is approximately 25,000 person-hours for about 300,000 incidents (12 incidents per hour).

In recent years, the AgInjuryNews collection, a publicly accessible repository of news articles and reports on agricultural injuries and fatalities, has been developed to address reporting gaps (Weichelt et al, 2018). Launched by the National Children’s Center for Rural Health and Safety (NCCRAHS) and curated by the National Farm Medicine Center (NFMC), AgInjuryNews has gathered over 4,500 incident reports involving more than 6,000 victims between 2015 and 2024 (AgInjuryNews, 2024). Data collection methods have been detailed elsewhere (Weichelt et al, 2018; Burke et al., 2021; Gorusu et al., 2020; Becklinger, 2024), but in brief, articles are sourced through a digital media service, screened, and summarized with key details and a link to the original report. Filters and keyword searches allow users to easily find relevant articles for research.

Over a dozen research articles based on this data have been published, with a full list available on the AgInjuryNews website (AgInjuryNews, 2024). The structured and searchable format of AgInjuryNews makes it a valuable resource for researchers and policymakers (AgInjuryNews, 2024). AgInjuryNews aims to provide an accessible resource for tracking trends, identifying emerging issues, promoting safety messaging, and raising awareness of agricultural injury prevention. However, the current scope of AgInjuryNews is limited. The process of identifying relevant articles and extracting necessary information for database entries is primarily manual and performed by volunteers, requiring a thorough understanding of the content. Developing a custom automated system to extract data from diverse news sources, especially one tailored to the specific format of each source, is neither economically nor technically feasible.

Predicting agriculture related injury severity is a crucial and promising area of research in safety studies. Gaining a deeper understanding of the factors that contribute to injury severity is essential for proactively implementing appropriate mitigation strategies. The severity of injuries is influenced by a combination of human factors, environmental conditions, roadway characteristics, and some special situations (Jamal et al., 2021). However, agriculture related injuries are random events that exhibit spatial variability, meaning the factors influencing severity can differ from one location to another. As a result, a localized analysis of risk factors is necessary for accurate crash prediction and the effective implementation of appropriate countermeasures.

In the literature, different statistical regression models have been widely used to model crash injury severity. Crash severity modeling often involves discrete outcome models such as binary, multinomial logit, and probit models, which have been widely used due to the discrete nature of crash severity levels (Azimi et al., 2020; Rifaat & Chin, 2007). More advanced models have emerged to address issues of heterogeneity and correlation, including Bayesian hierarchical (Huang et al., 2008), ordered logit (Azimi et al., 2020), multivariate (Aguero-Valverde & Jovanis, 2009), and random parameter models (Milton et al., 2008). Applications of logit, probit, and random parameter models have shown promise in analyzing injury severity based on various factors such as driver demographics, crash conditions, and vehicle types (Garrido et al., 2014; Kockelman & Kweon, 2002). Recently, Bayesian hierarchical models have also been used to explore factors like signalized intersections and camera presence in crash injury severity (Huang et al., 2008; Haque et al., 2010).

While these models offer clear mathematical interpretation and insight into individual predictor variables, they have several limitations. They often rely on assumptions about data distribution and linear relationships between variables, which, if violated, lead to biased results (Li et al., 2012; Wang & Kim, 2019; Zahid et al., 2020). Additionally, they tend to have poor prediction accuracy because of unobserved heterogeneity and multicollinearity within crash data (Savolainen & Mannering, 2007). To address these issues, more complex models are required, making them computationally intensive and difficult to implement (Savolainen et al., 2011). Alternatively, with recent advances in machine learning, new models have emerged to address these issues, though many still focus on prediction accuracy without enhancing understanding of severity risk factors. Feature-based analysis or explainable AI (XAI) methods are crucial for addressing this gap.

Machine learning (ML) methods have been increasingly used in crash severity modeling to address the limitations of traditional statistical models. Standard or linear ML models such as Artificial Neural Networks (ANN), Support Vector Machines (SVM), Naïve Bayes (NB) and Decision Trees (DT) have demonstrated their capability to model complex, nonlinear relationships between crash factors and injury outcomes. ANN models have shown superior predictive performance compared to statistical methods, particularly in studies using multi-layer perceptron (MLP) architectures. For example, Abdelwahab and Abdel-Aty (2001) reported that MLP outperformed ordered logit models with a classification accuracy of 65.6%. Other studies have applied ANN for crash severity prediction with notable success, including Kunt et al. (2012), who used ANN to predict crash outcomes on the Tehran-Ghom Freeway, achieving high performance based on R-values and error measures. SVM models, known for their ability to handle complex, high-dimensional data, have also been effective. Li et al. (2012) applied SVM to predict crash severity on Florida’s freeway diverge segments and found it superior to ordered logit models.

Similarly, Mokhtarimousavi et al. (2019) showed that SVM models, enhanced by optimization algorithms like particle swarm and harmony search, performed better than mixed logit models for work zone crashes. Naïve Bayes (NB) is a fast and effective machine learning classifier based on Bayes’ rule, assuming conditional independence between variables, which simplifies parameter estimation (Kakhki et al., 2019). Despite its simplicity, NB performs well in various classification tasks, particularly in binary classifications such as injury severity (Marucci-Wellman et al., 2017). Decision Trees (DT) have been popular in crash severity classification due to their simplicity and interpretability. Chong et al. (2004) and Abellán et al. (2013) used DT to classify crash injury severity levels, with results showing DT models outperforming ANN in some cases.

Ensemble ML models, particularly Random Forest (RF) and eXtreme Gradient Boosting (XGB), have gained significant attention in recent years for crash severity prediction due to their ability to combine multiple weak learners to improve predictive accuracy. RF, an ensemble of decision trees, has been widely used in crash severity studies because of its robustness and high performance. Zhang et al. (2018) compared RF with other machine learning and statistical models like SVM and ordered probit models, finding that RF consistently provided better predictions across different severity levels in freeway crashes. Similarly, Dadashova et al. (2020) demonstrated that RF outperformed traditional discrete choice models in predicting injury severity, identifying factors such as roadway design and crash type as crucial predictors.

XGB, a gradient boosting technique, has also emerged as a highly efficient and accurate model. Studies have shown that XGB surpasses other machine learning algorithms in both accuracy and computational efficiency. For instance, Parsa et al. (2020) applied XGB for real-time crash detection, achieving high detection rates and accuracy. Chen et al. (2020) used XGB to analyze crash severity in automated vehicle crashes and found it superior to classification and regression tree (CART) models. Similarly, Ma et al. (2019) reported that XGB, with a prediction accuracy of 86.73%, outperformed models like RF and SVM in predicting fatal crashes in Los Angeles, making it a powerful tool for road safety analysis. Despite their potential, these models face challenges such as the black-box nature of predictions, requiring explainable artificial intelligence (XAI) to enhance their interpretability and usability.

All the ML models previously discussed exhibit black-box characteristics. This means that the end user has access only to the input data and final predictions generated by the models (Saeed & Omlin, 2020; Sit et al., 2021). Consequently, users are often unaware of the reasoning behind the predictions made by complex AI systems and algorithms (Demiray et al., 2024). Recently, explainable artificial intelligence (XAI) has emerged to overcome the ‘black-box’ nature of ML models (Arrieta et al., 2020; Mermer et al., 2024). Among the various explanation techniques, Shapley additive explanation (SHAP) is the most representative post hoc analysis technique (Lundberg & Lee, 2017). SHAP can estimate the magnitude of the positive or negative contribution of the input features to a model’s output (Lundberg et al., 2020). Additionally, SHAP analysis is also utilized in accident injury severity prediction because it can effectively visualize the importance and effect of contributing factors on injury (Aboulola, 2024; Cicek et al., 2023; Yang et al., 2022).

The literature review above demonstrates that ML methods have been widely used for classifying and predicting future events across various fields, including occupational injury analysis. However, there is a gap in the literature regarding the evaluation of ML techniques for classifying and predicting the severity of occupational incidents in agriculture domain within the United States. Furthermore, to the best of our knowledge, there is no study to explore contributing factor with XAI and to compare different ML models’ performance to each other for agricultural injuries with US.

The primary goal of this study is to develop ensemble machine learning based injury severity prediction model and to identify major contributing factors utilizing an explainable AI approach. A comprehensive model comparison explores diverse models, enhancing understanding and highlighting ensemble ML adaptability. Specifically, our contributions are (1) utilizing extensive datasets that include various parameters for agriculture injury; (2) conducting experiments on various ML models including NB, SVM, XGB, stacking and voting models; (3) prediction of agricultural injury severity using these models; and (4) Demonstrating the significance of features through the utilization of the SHAP value.

The paper is structured as follows: Section 2 outlines the methodology, detailing the data collection process, feature preprocessing, and the implementation of various linear and ensemble machine learning models. Section 3 presents the results and discussion, including model performance evaluations, SHAP-based feature importance analysis, and interpretability insights. Section 4 concludes with a summary of key findings, practical implications for agricultural safety, and recommendations for future research directions.

## 2. Methodology

### 2.1. Case Study Dataset

The data used for this study was scraped from web by AgInjuryNews.com (Weichelt et al., 2018; AgInjuryNews, 2024). The website contains agricultural related injury incidents between 2015 and 2024 that occurred in US and Canada. For each incident, information about the location, data, age, as well as other factors related to the person during the time of the incident, such as if alcohol or any other drugs were present. The retrieved data was cleaned and preprocessed by deleting outliners, duplicate records, and records with missing information. The final dataset had a total of 2421 valid incident records that contained 757 non-fatal injuries and 1664 fatalities. The dataset had five explanatory categories with 30 child features (sub-levels) for categorical variables and two for continuous numerical variables.

Temporal characteristics include the time of the accident (morning, afternoon, evening or night hours), in which the incidents occurred. Variables under roadway or environmental characteristics comprised of location of the accident, confined space, grain involved, and agritourism. The explanatory variables under accident attribute includes injury agent where the accident occurred. The personal attribute consisted of gender, age, and role (part the occupant played in the incident), and state. Likewise, variables related to safety include alcohol, seatbelt, and helmet usage. The dependent variable in this study was the severity outcome of the incident and was classified in two levels (non-fatal and fatal). In this case, two level of severity outcome can be defined as follow: fatal that indicates that at least one person was killed during the incident and non-fatal includes property damage only and other minor injuries.

Table 1 summarizes the descriptive statistics of all the variables used in the machine learning study, offering insights into the temporal, environmental, accident-related, personal, and safety attributes of the incidents analyzed. The time-of-day variable is divided into four categories: morning (27.71%), afternoon (45.51%), evening (23.37%), and night (3.38%), with afternoon being the most frequent. In the roadway/environmental category, incidents mostly occurred on farmland or agricultural environments (49.58%) and roadways (46.74%), with a small percentage happening in forestry or fishing environments (3.68%). Only 6.26% of incidents involved confined spaces, and 6.76% involved grain, while agritourism was rare, with less than 1% occurrence each. The injury agent variable indicates that vehicles were the most common cause of incidents (61.62%), followed by machinery (10.94%) and ATVs/off-road vehicles (10.49%). For the personal category, males accounted for the vast majority of incidents (86.74%), with a mean age of 50. In terms of safety, very few incidents involved alcohol (1.69%) or the use of seatbelts (2.15%), or helmets (0.53%).

**Table 1.** Descriptive statistics of all variables in the dataset.

| Attribute | Data Description | Min | Max | Mean | Std. Dev | Percent |
| --- | --- | --- | --- | --- | --- | --- |
| **Temporal** |  |  |  |  |  |  |
| Time | 1: Morning (6am – 12pm) | 1 | 4 | 2.024 | 0.803 | 27.71 |
|  | 2: Afternoon (12pm – 6pm) |  |  |  |  | 45.51 |
|  | 3: Evening (6pm – 12am) |  |  |  |  | 23.37 |
|  | 4: Night (12am – 6am) |  |  |  |  | 3.38 |
| **Roadway / Environmental** |  |  |  |  |  |  |
| Location | 1: On Farmland or Ag. Environments | 1 | 3 | 1.972 | 0.981 | 49.58 |
|  | 2: Forestry or Fishing Environments |  |  |  |  | 3.68 |
|  | 3: Roadways |  |  |  |  | 46.74 |
| Confined Space | 0: No | 0 | 1 | 0.001 | 0.025 | 93.74 |
|  | 1: Yes |  |  |  |  | 6.26 |
| Grain Involved | 0: No | 0 | 1 | 0.068 | 0.250 | 93.24 |
|  | 1: Yes |  |  |  |  | 6.76 |
| Agri-tourism | 0: No | 0 | 1 | 0.008 | 0.087 | 99.24 |
|  | 1: Yes |  |  |  |  | 0.76 |
| **Accident** |  |  |  |  |  |  |
| Injury Agent | 1: ATV/Off-Road Vehicle | 1 | 9 | 7.342 | 2.717 | 10.49 |
|  | 2: Building |  |  |  |  | 2.60 |
|  | 3: Environment |  |  |  |  | 1.69 |
|  | 4: Fall |  |  |  |  | 1.82 |
|  | 5: Fishing/Forestry |  |  |  |  | 2.10 |
|  | 6: Livestock |  |  |  |  | 2.68 |
|  | 7: Machinery |  |  |  |  | 10.94 |
|  | 8: Pesticides/Plants |  |  |  |  | 6.03 |
|  | 9: Vehicle |  |  |  |  | 61.62 |
| **Personal** |  |  |  |  |  |  |
| Gender | 1: Male | 1 | 2 | 1.132 | 0.339 | 86.74 |
|  | 2: Female |  |  |  |  | 13.26 |
| Age | [0, 98] | 0 | 98 | 50.351 | 22.169 | ----- |
| State | [1, 59] | 1 | 59 | 30.558 | 15.465 | ----- |
| **Safety** |  |  |  |  |  |  |
| Alcohol | 0: No | 0 | 1 | 0.016 | 0.129 | 98.30 |
|  | 1: Yes |  |  |  |  | 1.69 |
| Seatbelt | 0: No | 0 | 1 | 0.021 | 0.145 | 97.85 |
|  | 1: Yes |  |  |  |  | 2.15 |
| Helmet | 0: No | 0 | 1 | 0.0054 | 0.073 | 99.46 |
|  | 1: Yes |  |  |  |  | 0.53 |

Table 2 provides the frequency and percentage distribution of injury severity categories (non-fatal and fatal) across different years from 2016 to 2024, highlighting a clear trend of decreasing injury frequencies over time. Out of the total 2,421 incident, 757 (31%) are classified as non-fatal injuries, 1,664 (69%) as fatal injuries. Fatal injuries consistently made up the majority of incidents, peaking in 2018 with 110 cases. However, from 2019 onwards, both fatal and non-fatal injuries begin to decline, with a notable shift in 2020 where non-fatal injuries rise to 33% of the total despite fewer overall incidents. By 2023 and into 2024, the frequency of both categories drops dramatically, especially in 2024, where only 11 non-fatal injuries (31%) and 24 fatal injuries (69%) are recorded, covering the period until May. This overall decline reflects a significant reduction in injury severity over the years, particularly in the most recent data.

**Table 2.** Frequency and percentage distribution of injury severity categories over the years.

| Year | Injury Severity Category | Frequency | Percent (%) |
| --- | --- | --- | --- |
| 2016 | Non-Fatal | 97 | 30% |
|  | Fatal | 225 | 70% |
| 2017 | Non-Fatal | 107 | 32% |
|  | Fatal | 225 | 68% |
| 2018 | Non-Fatal | 110 | 31% |
|  | Fatal | 248 | 69% |
| 2019 | Non-Fatal | 97 | 29% |
|  | Fatal | 246 | 72% |
| 2020 | Non-Fatal | 105 | 33% |
|  | Fatal | 212 | 67% |
| 2021 | Non-Fatal | 76 | 31% |
|  | Fatal | 170 | 69% |
| 2022 | Non-Fatal | 76 | 30% |
|  | Fatal | 181 | 70% |
| 2023 | Non-Fatal | 78 | 37% |
|  | Fatal | 133 | 63% |
| 2024 | Non-Fatal | 11 | 31% |
|  | Fatal | 24 | 69% |

### 2.2. Machine Learning Models

This section reviews the linear and ensemble machine learning classification algorithms and discusses their application for agricultural injury severity prediction. Machine learning classifiers are supervised training algorithms used in classifying datasets that can produce promising results due to their multi-dimensional data processing capability, flexibility in implementation, versatility, and superior predictive capabilities (Sit et al., 2020; Zhang et al., 2023). In this study, the target (dependent) variable (injury severity level) has two possible outcomes (non-fatal, and fatal). All the algorithms for current study were implemented using Python. The detailed methodology is further discussed in the following sections.

#### Naïve Bayes (NB)

The Naive Bayes algorithm is a widely used classifier known for its simplicity and efficiency in building probabilistic models (Hastie et al., 2009). It is based on Bayes’ Theorem, which calculates conditional probabilities to predict outcomes. As a supervised classification technique, it assumes strong independence among features, meaning that the presence of one feature in a class is independent of any other feature. Despite its high bias, Naive Bayes performs well even with small datasets due to its low variance. The algorithm predicts the probability that a data point belongs to a class, and the class with the highest posterior probability is selected as the predicted outcome. Bayesian classification provides practical learning algorithms and prior knowledge for observed data. This algorithm has been applied for crash severity prediction successfully (Sarkar et al., 2020; Arhin & Gatiba, 2020) and has success when dealing with classification tasks.

#### Decision Tree (DT)

Decision trees are a popular machine learning algorithm which can be used in a variety of classification-based tasks (Quinlan, 1986; Xu et al., 2019; Alabbad et al., 2022). Each tree contains multiple nodes and edges, where nodes represent different “questions”, while edges represent the answers to those questions. Any path taken using any combination of edges is a possible result produced by the decision tree. During training, decision trees are designed to minimize loss and error by weighing different paths differently depending on the dataset. To optimize the training process, splitting and pruning are employed. Splitting divides the dataset into multiple smaller sections, helping models generalize better on unseen data. Pruning, which removes sections of the decision tree that do not improve classification accuracy reduces overfitting and reduces the size of decision trees. Decision trees have recently been applied in forming new ensemble algorithms (Moral-García et al., 2019) and other classification and regression tasks (Azhar et al., 2022; Chang & Wang, 2006) for analyzing traffic accident.

#### K Nearest Neighbors (KNN)

The KNN is another commonly applied machine learning method in classification tasks that uses different types of distance metrics to identify the K most similar points and predict outputs on new data (Guo et al., 2003; Cikmaz et al., 2024). It is important to identify the most optimal value of k to achieve the highest accuracy because a k too small could lead to inaccurate predictions, while a k too large can lead to the overfitting of the model. Then, when considering the k nearest neighbors and their respective output classes, a weighted average is performed to determine the result of that specific data point. The KNN has been used successfully in both classification and regression tasks in many fields, especially in crash injury severity prediction (Santos et al., 2022).

#### Multi-Layer Perceptron (MLP)

The multi-layer perceptron (MLP) is a variation of the artificial neural network. It consists of three layers: the input layer, the hidden layer, and the output layer. Each layer of the MLP contains multiple perceptrons, which perform simple logistic regressions and are the base for the calculations in the model. The hidden layer can also be represented as a fully connected dense network of multiple perceptrons. During training, the MLP constantly updates the weights in each perceptron through backpropagation and the calculation of error (Bayar et al., 2009; Li and Demir, 2024). Finally, activation functions are differentiable functions that can learn these weights through gradient descents. The MLP has also been widely applied in vehicle accident injuries (Zhang et al., 2022).

#### Stochaistic Gradient Descent (SGD)

The stochastic gradient descent model is an ML method that starts with a random set of parameters and iteratively updates these parameters until it finds the minimum value of a function. It generates a hyperplane to separate data from two different classes, and it updates its parameters iteratively. One key hyperparameter of the stochastic gradient descent classifier is the learning rate, which determines the step size along the negative gradient. Finally, regularization and data randomness techniques are employed to reduce overfitting. The SGD classifier has been used in tasks where a linear relationship is present (Candefjord et al., 2021).

#### Support Vector Machine (SVM)

The support vector machine (SVM) is a supervised ML method and is commonly used in classification and regression analysis due to its good classification accuracy, ability to handle non-linear data, and its reduced tendency to the over-fitting problem (Xiang et al., 2021; Han et al., 2014; Sit et al., 2023). The algorithm fits the most optimal hyperplane to characterize data into two separate groups, where the further apart two different data points of different classes are, the better the hyperplane. Moreover, an additional advantage of the SVM is that it is robust to outliers in data. Finally, another essential part of the SVM is the kernel function, where data can be transformed to be mapped differently to improve the probability of finding an accurate hyperplane. The method has been successful with high accuracies in a traffic accident detection/prediction (Li et al., 2012; Jamal et al., 2021).

### 2.3. Ensemble Learning Models

Ensemble learning is a concept in machine learning that involves the fusion of multiple weaker learners to form one stronger and more powerful model. This technique is especially important in injury control and safety science because it helps reduce uncertainty in models and improve redundancy and accuracy, but there still remains a lack of a comparative analysis of the capabilities of these models, especially in injury prevention and safety. In this study, ensemble ML models were examined under 3 main categories: bagging, boosting, and other (i.e., voting, and stacking).

#### 2.3.1. Bagging Classifiers Methods

##### Random Forest (RF)

Random Forest is a popular bagging ensemble method consisting of multiple decision trees (Islam et al., 2024). By combining several weak learners, Random Forest enhances model performance (Breiman, 2001). Bagging involves bootstrapping, where models are trained on different subsets of the data, and their predictions are aggregated. The algorithm’s diversity and robustness come from training each tree on a different subset of features. While RF performs well with continuous variables and large datasets, it can be sensitive to noisy data due to uneven distribution and requires sufficiently deep trees to perform well. It has been used in both classification and regression tasks, as well as in crash severity predictions (Zhang et al., 2018; Jamal et al., 2021).

##### Deep Forest (DF)

Developed by Zhou et al. (2019), Deep Forest (DF) is a novel method that uses a stacking ensemble technique within a cascade structure, integrating deep learning with ensemble methods. Layers of random forests replace neurons in a neural network-like structure, and a sliding window scans the raw features. Inspired by stacking ensemble techniques, DF emphasizes model diversity by training each learner on different parts of the dataset. Though DF has limited applications in safety and transportation, it shows promise, especially in fields like injury/crash severity prediction.

#### 2.3.2. Boosting Classifiers Methods

##### Gradient Boosting (GB)

Gradient Boosting is another decision tree-based ensemble technique that uses boosting (Friedman, 2001). In boosting, each new model corrects errors made by its predecessor, with trees built sequentially. The model adjusts weights and minimizes loss through gradient descent. GB is widely used for model fusion and improving overall performance for severity prediction of car or motor crashes (Jeong, et al., 2018).

##### Extreme Gradient Boosting (XGBoost)

XGBoost is an enhancement of gradient boosting that includes regularization penalties to reduce overfitting (Chen & Guestrin, 2016). Like gradient boosting, XGBoost adds learners iteratively, focusing on errors made by previous learners. Its primary improvement lies in its regularization techniques, which boost generalization and scalability. XGBoost is highly efficient with large datasets and has been used in diverse fields (Sit et al., 2022), from pedestrian injury severity prediction (Wu et al., 2024) to railroad accident analysis (Bridgelall & Tolliver, 2021).

##### Light Gradient Boosting (LightGBM)

LightGBM is another popular boosting method that speeds up training by binning data points (Ke et al., 2017). Unlike traditional gradient boosting, LightGBM builds its tree leaf-wise rather than depth-wise, improving efficiency and reducing loss. Its use of gradient one-sided sampling allows it to focus on high-error data points, making it faster than XGBoost and effective in tasks requiring high efficiency (Dong et al.,2022).

##### Adaptive Boosting (Adaboost)

Adaboost combines several weak learners into a strong learner by adjusting weights based on their accuracy. Weak learners are trained on weighted versions of the data, with new weights assigned after each iteration to focus on incorrect predictions. Adaboost has been applied in a variety of fields, including accident severity prediction (Kashifi, & Ahmad, 2020; Li et al., 2023).

#### 2.3.3. Other Classifier Methods

##### Voting Classifier

The voting classifier is an ensemble learning technique used for classification tasks that combines the predictions of multiple individual classifiers to improve accuracy (Bauer and Kohavi, 1999). Unlike bagging and boosting ensemble methods, the Voting Classifier simply aggregates predictions from several models that are trained independently and then combines their results. There are two main types of voting used in the voting classifier: hard voting and soft voting. Hard voting is simply a majority vote, where the most common class among the base estimators is the final predicted class. Soft voting outputs the probabilities for each class, and these probabilities are averaged to determine the weight of each class. The voting classifier aggregates the strengths of multiple models, which prevents the weaknesses of a single model from impeding accuracy significantly (Fahad et al., 2023).

##### Stacking Classifier

The stacking classifier is another ensemble learning technique that combines multiple different models to improve performance by taking advantage of their individual strengths (Wolpert, 1992). Unlike the voting classifier, which directly averages the predictions of each model, the stacking classifier uses another model to make the final prediction. This model is known as the final estimator or the meta-learner, and it optimally reduces prediction errors. The meta-learner can learn to integrate the strengths of each model, and it compensates for their individual model weaknesses. This allows the stacking classifier to improve accuracy and is powerful in situations with difficult datasets to analyze (Tang et al., 2019).

### 2.4. Model Development

Figure 1 shows a comprehensive workflow diagram for ML agricultural injury severity prediction. Initial step involves obtaining and preparing data from AgInjuryNews. After applying the preprocessing, organized data are split into training and test dataset, where the chosen ML models are trained using the training dataset, and test dataset are employed to observe the behavior of the trained ML models. Since our dataset contains many more samples than features, we were careful not to reduce the number of features too much to avoid losing important information. This strategy helps us balance capturing a wide range of influencing factors and maintaining the robustness of our model. The min-max method (MinMaxScaler) was applied to standardize the features to eliminate potential model bias due to difference in scales and units (Ahsan et al. 2021). The min-max method transforms the data into a range 0 and 1 and ensures that all features contribute equally to the model by bringing them to a common scale, which helps improve the performance of machine learning algorithms.

**Figure 1.**
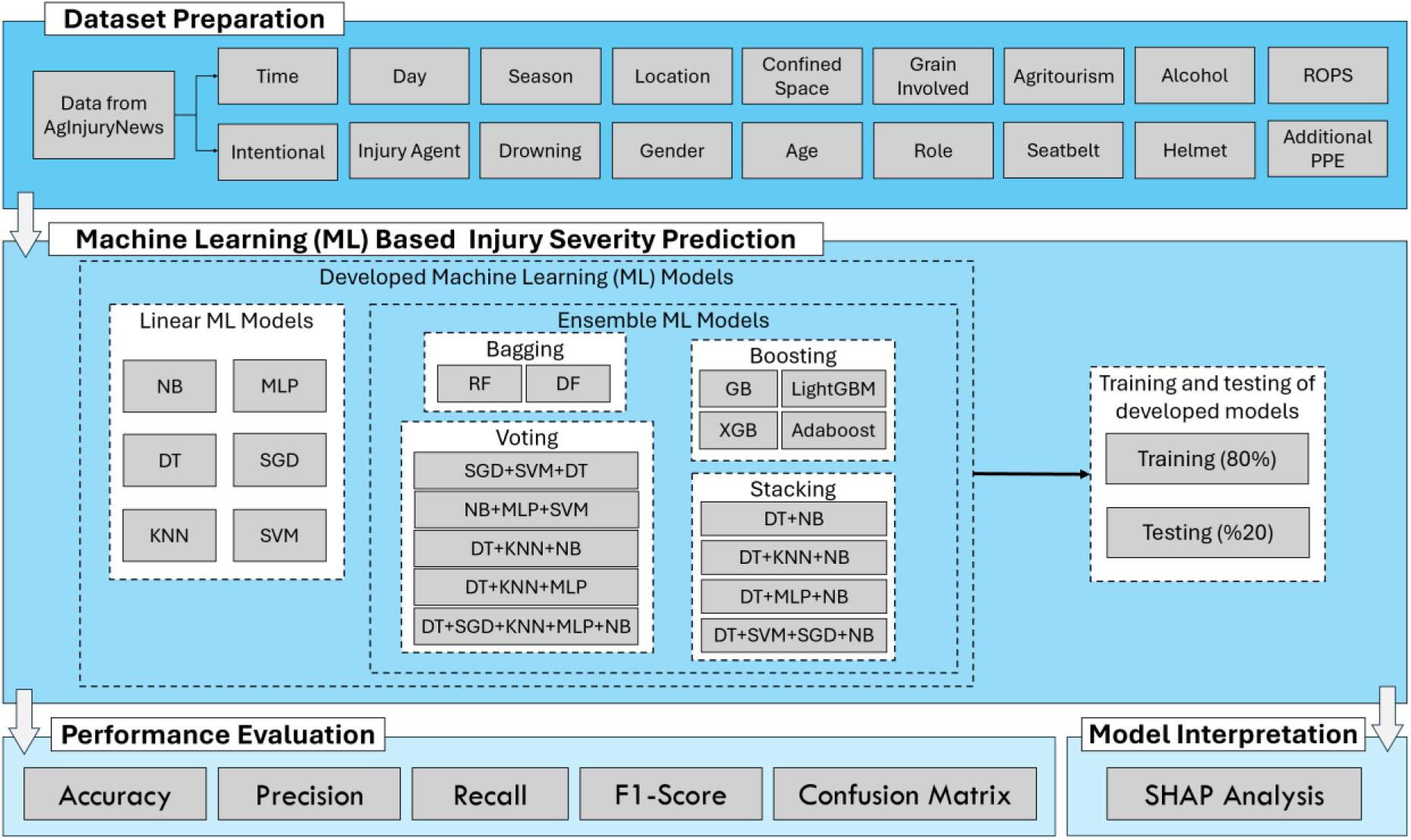
Workflow diagram for all scenarios in agricultural injury severity prediction.

A variety of ML models are then trained and tested to predict agricultural injury severity. Table 3 lists the training parameters for each of the selected ML models used in this study. The tuning of these hyperparameters was achieved through cross validation, where the performance of models under different parameters would be evaluated until the most accurate model setting was determined. Various combinations of individual ML models were evaluated for ensemble modeling. For the voting and stacking classifiers, different combinations of two to five linear ML models were tested. These models included NB, SVM, MLP, KNN, SGD, and DT, as they all achieved R² values above 0.5 but showed significant variation among themselves. Therefore, different combinations of these models were explored to assess the impact on accuracy when combining stronger and weaker learners.

**Table 3.** Summary of training parameters of models developed in this study.

| **Models** | **Parameters** |
| --- | --- |
| NB | alpha = 21.62 |
| KNN | n_neighbors = 47, leaf_size = 38 |
| DT | min_samples_split = 43, min_samples_leaf = 36, max_depth = 5 |
| SGD | max_iter = 1129, tol = 0.602, alpha = 3.076 |
| MLP | Max_iter = 1000, tol = 1e-4 |
| SVM | C = 3.846, tol=0.601, degree = 1732 |
| RF | Max_depth = 100, max_feature_split = 0.1, max_sample_split = 50.0, n_estimator = 250 |
| DF | N_estimators = 10 |
| GB | Learning_rate = 0.2526, max_depth 1, max_feature_split = 83, min_samples_split = 33, n_estimators = 173.0 |
| LightGBM | Learning_rate = 0.1303, max_depth = 54, n_estimators = 16, num_leaves= 2 |
| XGBoost | Colsample = 1.0, gamma = 4.264, learning_rate = 0.5384, max_depth = 4, n_estimators = 240 |
| AdaBoost | Learning_rate = 0.05818, n_estimators = 217 |

### 2.5. Model Evaluation

When comparing classification models, various performance metrics derived from a confusion matrix are commonly used. Table 4 presents the confusion matrix for a binary classifier. This matrix, structured as a contingency table, illustrates how observations are distributed across actual and predicted classes (Guns et al., 2012). In classification problems, the confusion matrix comprises four possible outcomes: true positives (TP), true negatives (TN), false positives (FP), and false negatives (FN), as shown in Table 3. It provides a clear representation of both correct and incorrect classifications under a specified target, allowing for the calculation of accuracy and other performance metrics.

**Table 4.** Confusion Matrix for Binary Classification.

| Actual Class | Predicted Class |  |
| --- | --- | --- |
|  | Positive | Negative |
| Positive | True Positive (TP) | False Negative (FN) |
| Negative | False Positive (FP) | True Negative (TN) |

In this study, the binary confusion matrix for each machine learning model was utilized to compute various quantitative performance metrics. The metrics used for evaluating model performance are outlined and explained below (Sokolova and Lapalme, 2009; Guns et al., 2012; Mathew, 2016; Shreve et al., 2011). Accuracy, expressed by Equation 1, represents the proportion of correctly classified samples out of the total number of samples. Precision (equation 2) assesses the alignment between data labels and the positive labels predicted by the classifier. Recall, or sensitivity (equation 3), indicates how effectively the classifier identifies positive labels. Finally, the F1-score (equation 4) is the harmonic mean of recall and precision, providing a balanced measure of the that considers both false positives and false negatives in a classification model.

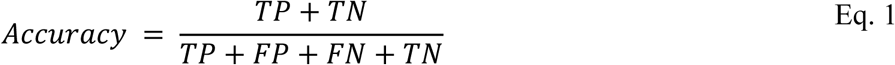

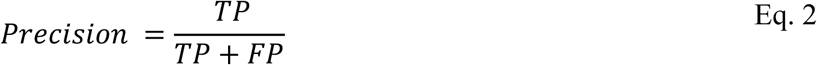

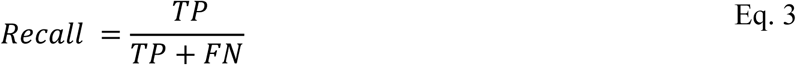

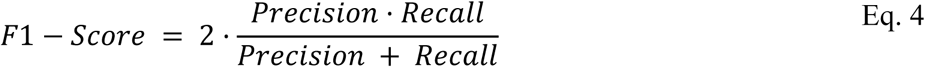

## 3. Results and Discussion

Figure 2 presents the distribution of injury severity, comparing fatal and non-fatal injuries over the years from 2016 to 2024, with the 2024 data covering the period until May. In 2016, the frequency of fatal injuries is relatively high, with around 225 incidents. This trend continues to rise, peaking in 2018 when the number of fatal injuries reaches nearly 250. However, after 2019, there is a noticeable decline in fatal injuries. By 2023 and the early months of 2024, the frequency of fatal injuries sharply decreases. On the other hand, non-fatal injuries exhibit a more stable pattern from 2016 to 2020. In 2018, non-fatal injuries peak slightly above 110, before beginning to decline. The reduction becomes more noticeable after 2021, following a downward trajectory similar to fatal injuries.

**Figure 2.**
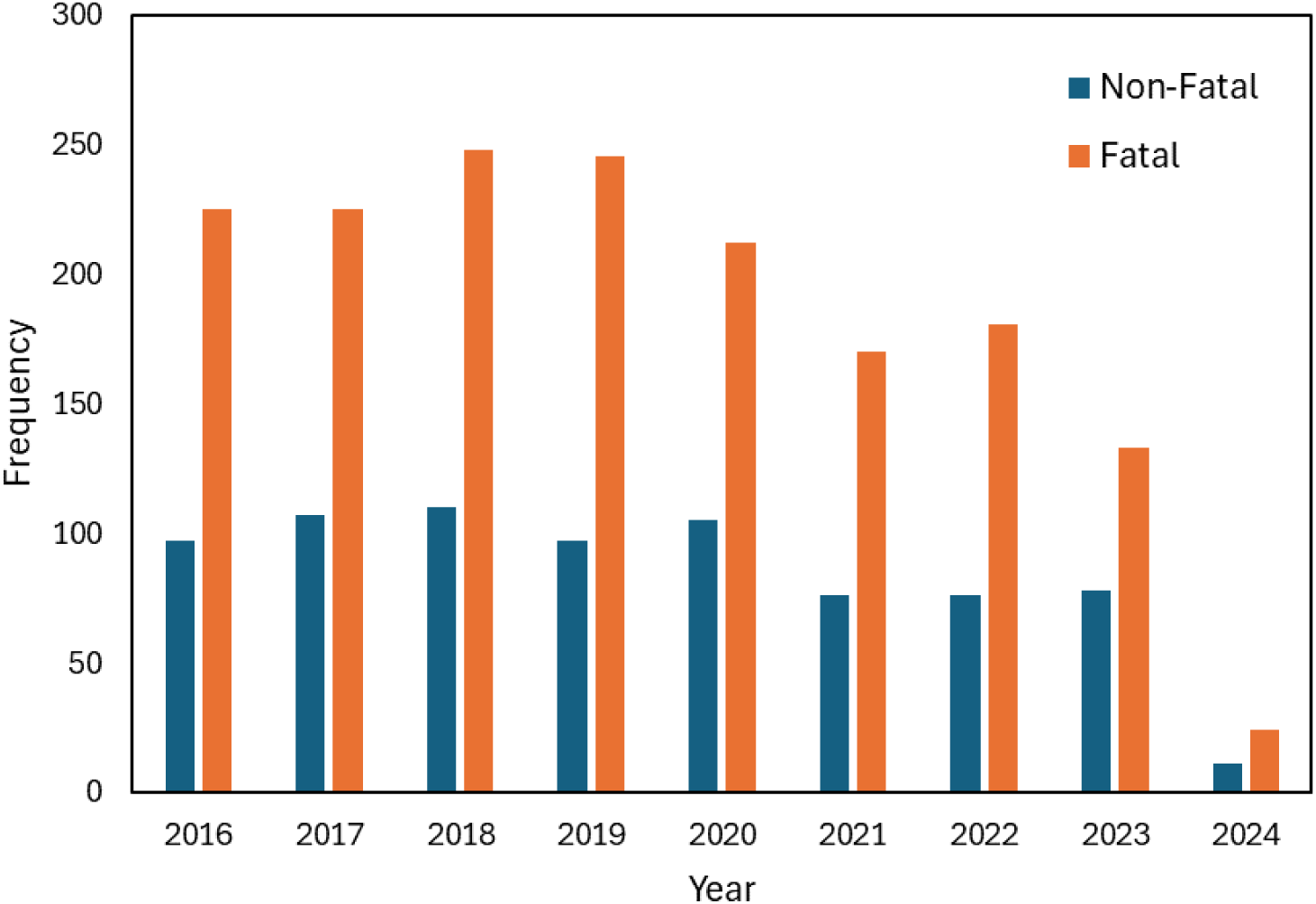
The distribution of injury severity categories over the years from 2016 to 2024

By 2023, and up until May 2024, the frequency of non-fatal injuries has drastically decreased. The trends in both fatal and non-fatal injuries suggest a significant improvement in safety measures or preventive strategies after 2020. The earlier and more gradual decline in fatal injuries begins in 2019, while non-fatal injuries show a sharper decrease starting in 2021. The data for 2024, though incomplete, already indicates a continuation of this downward trend, with both fatal and non-fatal injury frequencies approaching zero. This suggests that, by mid-2024, substantial progress has been made in reducing injury rates, likely due to enhanced interventions or safety regulations.

Table 5 presents the confusion matrices for six linear machine learning models—NB, DT, KNN, MLP, SGD, and SVM—used to predict injury severity in agriculture. The matrices provide insight into the models’ abilities to classify injuries as ’Fatal’ or ’Non-Fatal’, highlighting varying levels of accuracy across the models. The NB model shows a pronounced tendency to classify most cases as ’Fatal’, resulting in a high sensitivity (98.82% correct classification of actual ’Fatal’ cases) but poor specificity, as evidenced by the 94.44% misclassification of ’Non-Fatal’ cases.

**Table 5.** Confusion Matrix for All Linear ML Methods.

| Model | Actual Class | Predicted Class |  |
| --- | --- | --- | --- |
|  |  | Non-Fatal | Fatal |
| NB | Non-Fatal | 8 (5.56%) | 136 (94.44%) |
|  | Fatal | 4 (1.18%) | 337 (98.82%) |
| DT | Non-Fatal | 38 (26.39%) | 106 (73.61%) |
|  | Fatal | 41 (12.02%) | 300 (87.97%) |
| KNN | Non-Fatal | 20 (13.89%) | 124 (86.11%) |
|  | Fatal | 17 (4.98%) | 324 (95.02%) |
| MLP | Non-Fatal | 18 (12.50%) | 126 (87.50%) |
|  | Fatal | 19 (5.57%) | 322 (94.43%) |
| SGD | Non-Fatal | 0 (0.00%) | 144 (100.00%) |
|  | Fatal | 0 (0.00%) | 341 (100.00%) |
| SVM | Non-Fatal | 19 (13.19%) | 125 (86.81%) |
|  | Fatal | 17 (4.98%) | 324 (95.02%) |

In contrast, DT and KNN provide more balanced results, though they still misclassify a significant number of ’Non-Fatal’ cases as ’Fatal’. The MLP model performs similarly to KNN, displaying moderate classification capabilities for both categories. Notably, SGD exhibits severe bias, classifying all cases as ’Fatal’, leading to zero specificity and raising concerns about its utility in applications where identifying ’Non-Fatal’ cases is crucial. SVM offers a balanced performance, akin to KNN and MLP, with a reasonable trade-off between sensitivity and specificity.

This lower performance for ’Non-Fatal’ predictions can largely be attributed to the issue of data imbalance, which biases model estimations toward the majority class. This finding aligns with intuition and is consistent with observations reported in the literature (Jamal et al., 2021; Jiang et al., 2022). In this dataset, ’Fatal’ injuries likely constitute the majority class, leading the models to skew their predictions toward this category. As a result, the models provide biased estimations that favor the majority class, sacrificing specificity in identifying ’Non-Fatal’ injuries. This imbalance impacts the models’ ability to generalize effectively across both categories, underscoring the importance of addressing class distribution in training data to enhance model performance, particularly for the minority class.

In Table 6, the performance of linear machine learning models is evaluated using key metrics such as accuracy, precision, recall, and F1-score. These metrics provide a comprehensive assessment of each model’s ability to predict injury severity in agriculture. Accuracy measures the overall correctness of predictions, precision focuses on the model’s ability to minimize false positives, recall highlights the model’s sensitivity to true positives, and F1-score balances precision and recall, particularly useful when dealing with imbalanced datasets. Together, these metrics illustrate each model’s strengths and trade-offs between identifying true positives and avoiding false positives.

**Table 6.** Performance Metrics for all Linear ML Models.

| Model | Accuracy | Precision | Recall | F1-Score |
| --- | --- | --- | --- | --- |
| NB | 0.711 | 0.698 | 0.988 | 0.613 |
| DT | 0.697 | 0.662 | 0.879 | 0.660 |
| KNN | 0.711 | 0.674 | 0.953 | 0.644 |
| MLP | 0.701 | 0.650 | 0.944 | 0.633 |
| SGD | 0.703 | 0.494 | 1.000 | 0.580 |
| SVM | 0.707 | 0.664 | 0.950 | 0.639 |

Comparing the models, the NB model shows the highest recall (99%), making it highly sensitive to positive cases, though its precision (70%) and F1-score (61%) reflect some trade-off in false positives. The DT model performs the weakest overall, with lower metrics across the board, including an accuracy of 69.7% and F1-score of 66%. KNN achieves a strong balance with high recall (95%) and accuracy (71%), though its precision (67%) is slightly lower. MLP model performs similarly to KNN, with strong recall (94%) but slightly lower precision (65%) and F1-score (63%).

The SGD model, despite its perfect Recall (100%), suffers from very low precision (49%), indicating many false positives and a lower F1-score (58%). Lastly, the SVM offers a good overall balance with high recall (95%), moderate precision (66%), and an accuracy of 70.7%, making it one of the better-performing models in the table. In summary, KNN and SVM offer a balanced performance, while models such as NB and SGD excel in recall but face challenges with precision. This comparative evaluation provides valuable insights into each model’s predictive capabilities and highlights the trade-offs between sensitivity and precision in predicting injury severity in agricultural settings.

Table 7 presents the confusion matrices for various ensemble machine learning models applied to predict injury severity in agriculture, categorized into Non-Fatal and Fatal outcomes. The table highlights the classification performance of Bagging, Boosting, Stacking, and Voting ensemble methods by summarizing the true positives (TP), true negatives (TN), false positives (FP), and false negatives (FN) for each model. Across all models, there is a noticeable trend of high accuracy in predicting Fatal cases, with most models achieving TP rates exceeding 93%. However, challenges persist in correctly identifying Non-Fatal cases, as indicated by the consistently high FN rates, ranging from 81.25% to 100% for certain configurations.

**Table 7.** Confusion Matrix for All Ensemble ML Models.

| Ensemble Model |  | Actual Class | Predicted Class |  |
| --- | --- | --- | --- | --- |
|  |  |  | Non-Fatal | Fatal |
| Bagging | RF | Non-Fatal | 19 (13.19%) | 125 (86.81%) |
|  |  | Fatal | 11 (3.23%) | 330 (96.77%) |
|  | DF | Non-Fatal | 23 (15.97%) | 121 (84.03%) |
|  |  | Fatal | 18 (5.28%) | 323 (94.72%) |
| Boosting | GB | Non-Fatal | 24 (16.67%) | 120 (83.33%) |
|  |  | Fatal | 17 (4.99%) | 324 (95.01%) |
|  | XGB | Non-Fatal | 16 (11.11%) | 128 (88.89%) |
|  |  | Fatal | 12 (3.52%) | 329 (96.48%) |
|  | Adaboost | Non-Fatal | 18 (12.50%) | 126 (87.50%) |
|  |  | Fatal | 13 (3.81%) | 328 (96.19%) |
|  | LightGBM | Non-Fatal | 0 (0.00%) | 144 (100%) |
|  |  | Fatal | 0 (0.00%) | 341 (100%) |
| Stacking | S#1 (DT+KNN+NB) | Non-Fatal | 25 (17.36%) | 119 (82.64%) |
|  |  | Fatal | 22 (6.45%) | 319 (93.55%) |
|  | S#2 (DT+MLP+NB) | Non-Fatal | 11 (7.64%) | 133 (92.36%) |
|  |  | Fatal | 16 (4.69%) | 325 (95.31%) |
|  | S#3 (DT+NB) | Non-Fatal | 7 (4.86%) | 137 (95.14%) |
|  |  | Fatal | 4 (1.17%) | 337 (98.83%) |
|  | S#4 (DT+SVM+SGD+NB) | Non-Fatal | 27 (18.75%) | 117 (81.25%) |
|  |  | Fatal | 21 (6.16%) | 320 (93.84%) |
| Voting | V#1 (DT+KNN+MLP+SGD+NB) | Non-Fatal | 12 (8.33%) | 132 (91.67%) |
|  |  | Fatal | 10 (2.93%) | 331 (97.07%) |
|  | V#2 (DT+KNN+MLP) | Non-Fatal | 16 (11.11%) | 128 (88.89%) |
|  |  | Fatal | 13 (3.81%) | 328 (96.19%) |
|  | V#3 (DT+KNN+NB) | Non-Fatal | 15 (10.42%) | 129 (89.58%) |
|  |  | Fatal | 11 (3.23%) | 330 (96.72%) |
|  | V#4 (NB+MLP+SVM) | Non-Fatal | 18 (12.50%) | 126 (87.50%) |
|  |  | Fatal | 17 (4.99%) | 324 (95.01%) |
|  | V#5 (SGD+SVM+DT) | Non-Fatal | 7 (4.86%) | 137 (95.14%) |
|  |  | Fatal | 4 (1.17%) | 337 (98.83%) |

The Bagging model, specifically Random Forest (RF), demonstrates robust performance in Fatal predictions, achieving a TP rate of 96.77%. However, it struggles with Non-Fatal predictions, misclassifying 86.81% of actual Non-Fatal cases as Fatal, indicating a strong bias towards the dominant class. Boosting models, including Gradient Boosting (GB), XGBoost (XGB), and AdaBoost, exhibit similar trends. Among these, XGB achieves the highest TP rate of 96.48% for Fatal cases, while GB demonstrates a slightly better balance with a lower FN rate for Non-Fatal predictions (83.33%). Conversely, LightGBM, although achieving 100% accuracy for Fatal cases, completely fails to classify Non-Fatal outcomes correctly, suggesting extreme sensitivity to the class imbalance in the dataset.

In this work, the stacking model was developed using DT algorithm as a meta learning model due to its ability to capture complex, non-linear relationships between diverse base learner predictions. Additionally, DTs are robust to noisy or conflicting base model predictions, providing more stable and accurate final classifications. Models with the highest accuracy score were chosen and paired with a DT and based on the results obtained we realized that combining a strong model with DT increases its accuracy value Stacking models combine predictions from multiple base models, yielding varied results based on the chosen configuration. The stacking model S#3 (DT+NB) stands out, achieving the highest TP rate for Fatal cases (98.83%) while also maintaining the lowest FN rate for Non-Fatal cases (95.14%). This indicates its superior ability to balance predictions across both classes. In contrast, more complex configurations, such as S#4 (DT+SVM+SGD+NB), although achieving high accuracy for Fatal cases (93.84%), show reduced performance for Non-Fatal predictions, with an FN rate of 81.25%.

Voting ensembles, which aggregate predictions through majority or weighted voting, show varied effectiveness depending on the model combination. V#5 (SGD+SVM+DT) and V#3 (DT+KNN+NB) demonstrate the most balanced performance, achieving TP rates of 98.83% and 96.72%, respectively, for Fatal cases while maintaining relatively low FN rates for Non-Fatal predictions. However, other configurations, such as V#1 (DT+KNN+MLP+SGD+NB), prioritize Fatal predictions, achieving a TP rate of 97.07% but at the expense of higher FN rates for Non-Fatal outcomes (91.67%). This suggests that ensemble learning methods, particularly boosting and voting strategies, in accurately identifying high-severity Fatal injuries. However, the elevated FN rates for Non-Fatal cases across most models point to the need for further optimization to address class imbalance. These findings highlight the potential of ensemble models while emphasizing the require further tuning to improve precision in critical agricultural injury prediction scenarios.

Table 8 shows a comprehensive analysis of performance metrics across various ensemble ML models, including bagging, boosting, stacking, and voting approaches. Among the bagging models, RF performs best with an accuracy of 72%, high recall (99%), and moderate precision (76%), reflecting its strong ability to identify Fatal cases. However, DF, while achieving a similar recall (97%), shows lower precision (67%), indicating a higher rate of false positives and reduced overall balance compared to RF. For boosting models, AdaBoost achieves the highest accuracy (71%) and F1-score (64%) in its category, demonstrating a good balance between precision (68%) and recall (96%).

**Table 8.** Performance Metrics for All Ensemble ML Models.

| **Ensemble Model** |  | **Accuracy** | **Precision** | **Recall** | **F1-Score** |
| --- | --- | --- | --- | --- | --- |
| Bagging | RF | 0.715 | 0.759 | 0.986 | 0.621 |
|  | DF | 0.707 | 0.665 | 0.973 | 0.619 |
| Boosting | GB | 0.709 | 0.670 | 0.933 | 0.655 |
|  | XGB | 0.714 | 0.477 | 1.000 | 0.565 |
|  | Adaboost | 0.713 | 0.680 | 0.962 | 0.641 |
|  | LightGBM | 0.703 | 0.494 | 1.000 | 0.580 |
| Stacking | S#1 (DT+KNN+NB) | 0.709 | 0.669 | 0.935 | 0.653 |
|  | S#2 (DT+MLP+NB) | 0.692 | 0.619 | 0.953 | 0.610 |
|  | S#3 (DT+NB) | 0.709 | 0.688 | 0.988 | 0.608 |
|  | S#4 (DT+SVM+SGD+NB) | 0.715 | 0.681 | 0.938 | 0.661 |
| Voting | V#1 (DT+KNN+MLP+SGD+NB) | 0.707 | 0.664 | 0.970 | 0.621 |
|  | V#2 (DT+KNN+MLP) | 0.709 | 0.669 | 0.961 | 0.633 |
|  | V#3 (DT+KNN+NB) | 0.711 | 0.676 | 0.967 | 0.632 |
|  | V#4 (NB+MLP+SVM) | 0.705 | 0.658 | 0.950 | 0.635 |
|  | V#5 (SGD+SVM+DT) | 0.709 | 0.688 | 0.988 | 0.608 |

GB follows closely with comparable accuracy (71%) and a slightly higher F1-score (66%), although it exhibits slightly lower precision (67%). XGB and LightGBM exhibit perfect recall (100%), excelling in identifying Fatal cases. However, their precision values (48% and 49%, respectively) are significantly lower, resulting in compromised F1-scores of 57% and 58%. These results highlight that while XGB and LightGBM are sensitive to Fatal cases, they frequently misclassify non-fatal cases, reflecting an imbalance in their predictions.

Stacking ensemble models present more varied performance. Stacking configuration S#4 achieves the highest accuracy (71.5%) and F1-score (66%) in this category, with balanced precision (68%) and recall (94%), making it the most robust stacking approach. S#3 shows the highest recall (99%) among stacking models, indicating strong sensitivity to Fatal outcomes, but its moderate precision (69%) results in a lower F1-score (61%). S#2 delivers the weakest performance with an accuracy of 69% and F1-score of 61%, indicating limited predictive capability in this configuration.

Voting ensemble models demonstrate consistent performance, with accuracy ranging from 70% to 71%. Among these, V#3 and V#4 configurations achieve balanced precision (68% and 66%, respectively) and recall (98% and 95%), resulting in F1-scores of 63% and 64%. V#5 shows the highest recall (99%) but achieves only moderate precision (69%), leading to an F1-score of 61%. These results indicate that while voting ensembles effectively predict Fatal cases, their ability to correctly identify Non-Fatal cases vary depending on the model combination. This suggests that while ensemble ML models are highly effective in identifying Fatal injury outcomes, their performance in classifying Non-Fatal cases remains a challenge due to imbalanced precision and recall. This highlights the need for further optimization, such as incorporating cost-sensitive learning, class balancing techniques, or domain-specific feature engineering, to enhance their predictive balance. These findings emphasize the importance of selecting and fine-tuning ensemble configurations to achieve a more equitable balance between sensitivity and specificity in injury severity prediction.

Shapley Additive exPlanations (SHAP) values are used to decode the “black box” behind machine learning models by assigning interpretability to their predictions. Derived from game theory, SHAP values distribute a model’s prediction by evaluating each feature’s contribution to the outcome. This is achieved by calculating the average contribution of a feature across all possible combinations of inputs, including the impact of removing that feature. Through this method, we gain insight into the relative importance of input features in determining predictions.

In this study, we utilized the SHAP values to analyze the contributions of input features within various ML models for predicting agricultural injury severity. Both TreeSHAP and KernelSHAP can be used to estimate the SHAP values in each model. These functions allow us to generate bar plots and bee swarm plots of the effect of different features on the output for each data point. The relative importance values (i.e., the mean |SHAP| value) provide insights into the contribution of each feature to the model’s predictions, enabling us to understand the driving factors behind the predictions. Table 9 highlights the mean SHAP values for the top five features in each model, illustrating their relative importance. The higher the SHAP value for a feature, the more it influences the model’s prediction.

**Table 9.** SHAP Values for Individual Ensemble and Linear ML Models.

| ML Model | Input Features (mean\|SHAP\|) |  |  |  |  |
| --- | --- | --- | --- | --- | --- |
| **NB** | Seatbelt<br>(0.035) | Gender<br>(0.055) | Location<br>(0.008) | Injury Agent<br>(0.007) | Time<br>(0.05) |
| **DT** | Age<br>(0.141) | Location<br>(0.100) | Gender<br>(0.080) | Time<br>(0.070) | State<br>(0.370) |
| **KNN** | Location<br>(0.062) | Gender<br>(0.060) | Age<br>(0.050) | State<br>(0.045) | Time<br>(0.035) |
| **MLP** | Time<br>(0.065) | Age<br>(0.049) | Gender<br>(0.048) | Location<br>(0.045) | Seatbelt<br>(0.026) |
| **SGD** | Location<br>(0.014) | Agent<br>(0.006) | Gender<br>(0.0059) | State<br>(0.0025) | Age<br>(0.001) |
| **SVM** | Gender<br>(0.08) | State<br>(0.03) | Seatbelt<br>(0.027) | Time<br>(0.025) | Age<br>(0.024) |
| **RF** | Age<br>(0.135) | Location<br>(0.10) | Agent<br>(0.085) | State<br>(0.082) | Time<br>(0.080) |
| **DF** | Gender<br>(0.060) | Age<br>(0.048) | State<br>(0.036) | Time<br>(0.034) | Location<br>(0.032) |
| **GB** | Age<br>(0.25) | Gender<br>(0.14) | Time<br>(0.13) | Location<br>(0.11) | Agent<br>(0.10) |
| **XGB** | Age<br>(0.22) | Agent<br>(0.125) | Location<br>(0.117) | Gender<br>(0.100) | Time<br>(0.095) |
| **LGBM** | Age<br>(0.13) | Location<br>(0.078) | Gender<br>(0.072) | Time<br>(0.02) | Seatbelt<br>(0.019) |
| **Adaboost** | Gender<br>(0.042) | Seatbelt<br>(0.030) | Location<br>(0.027) | Age<br>(0.026) | Time<br>(0.017) |

The results indicate notable variations in feature importance across the evaluated models. NB emphasizes Gender and Time as the most significant predictors, highlighting the importance of demographic and temporal factors, while DT assigns significant weight to State, followed by Age, Location, Gender, suggesting the influence of geographical and demographic factors. The KNN model focuses on Location and Gender, emphasizing spatial and demographic features, while the MLP demonstrates a relatively balanced distribution, with Time, Age, and Gender as the top contributors.

Linear models like SGD and SVM show lower overall SHAP values, with SGD relying on Location and Agent and SVM emphasizing Gender and State. Tree-based ensemble models such as RF and DF place higher importance on Time, Age and Gender, reflecting their sensitivity to demographic and temporal factors. Boosting algorithms, including GB, XGB, and LightGBM, consistently prioritize Age and Gender, with GB and XGB assigning the highest SHAP values to these features. AdaBoost, while more evenly distributed, also emphasizes Gender and Seatbelt as the primary contributors.

Across all models, Age, Gender, Time, and Location emerge as consistent predictors of agricultural injury severity, underscoring their universal relevance. Ensemble models like GB and XGB exhibit the strongest emphasis on these features, while linear models like SGD show a weaker influence. The integration of SHAP values provides critical insights into the interpretability of these models, enabling a better understanding of how individual features drive predictions, which is essential for practical application in agricultural safety initiatives.

SHAP provides both the direction and magnitude of feature contributions to prediction results, offering a transparent way to interpret machine learning models. Figure 3 illustrates SHAP summary plots for the KNN and XGB models, showing the relative effect of each input feature on the models’ predictions. In these plots, the input features are sorted by their SHAP values, with the most impactful features appearing at the top. Each dot represents a SHAP value for a specific prediction, plotted along the x-axis to indicate the contribution of the feature to the model’s output. The color of the dots represents the actual observed values of the features, ranging from high (red) to low (blue).

**Figure 3.**
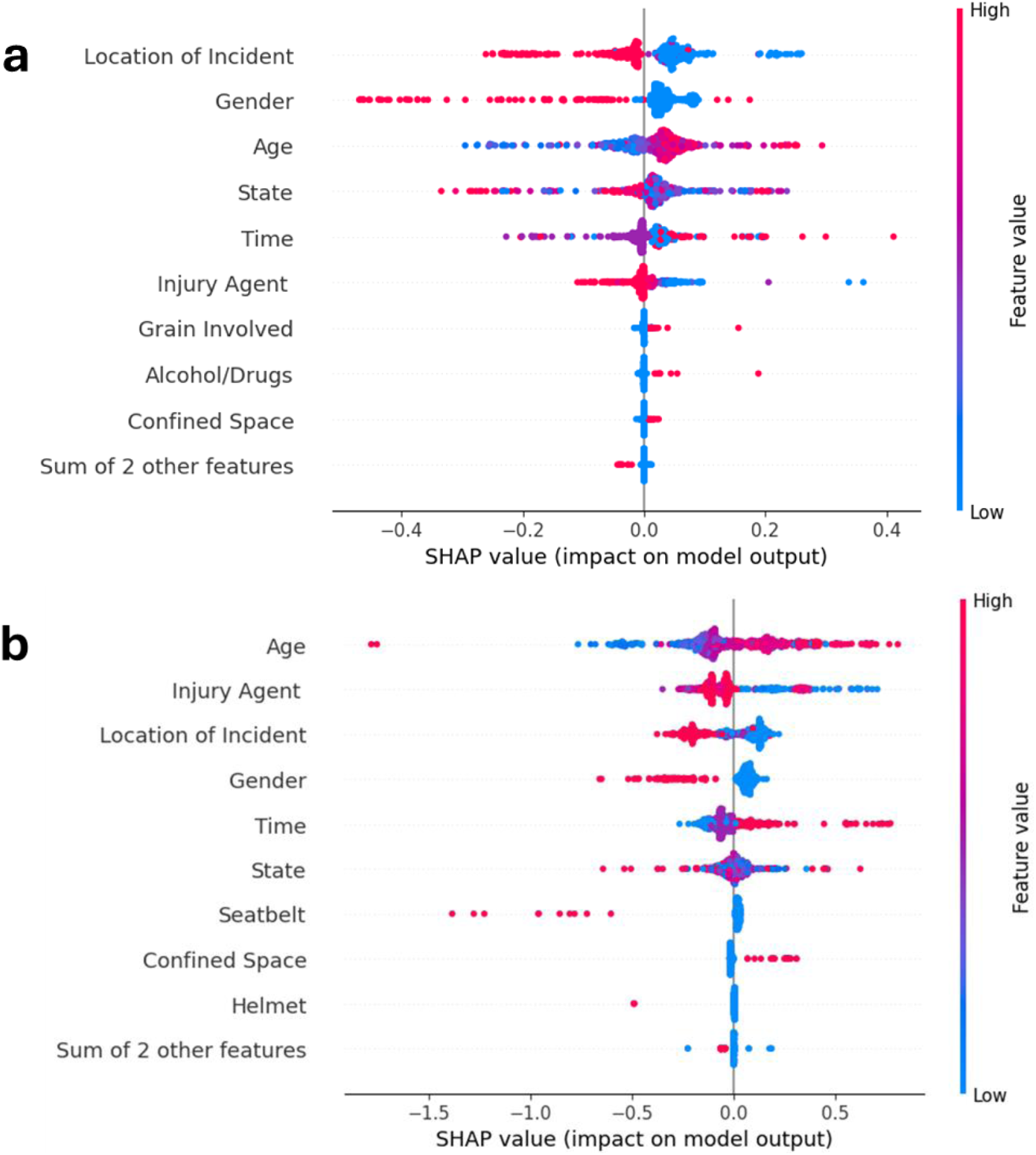
Beeswarm plot of SHAP value of features and impact on the model output for (a) KNN and (b) XGB

As shown in Figure 3, the Location of the incident emerges as the most influential feature in both models. Higher values of this feature, such as incidents occurring in roadways, result in lower SHAP values, corresponding to a negative impact on the model’s prediction. This aligns with the observation that rural and farmland environments are more prone to severe accidents due to unique risk factors associated with agricultural activities (Aboulola et al., 2024). The next significant feature is Gender, with lower values (indicating male individuals) contributing more strongly to predictions of severe injuries. This finding aligns with studies showing that male individuals are generally at higher risk due to behavioral factors such as increased risk-taking tendencies (Bener & Crundall, 2008).

Another critical feature is Age, which shows a positive correlation with injury severity. As age increases, the SHAP values indicate a stronger contribution to the likelihood of severe injuries, consistent with research suggesting that older individuals are more vulnerable to severe outcomes in accidents (Tavris et al., 2001). Overall, the SHAP summary plots provide an interpretable and actionable explanation of the KNN and XGB models, emphasizing the importance of demographic, environmental, and situational factors in predicting agricultural injury severity. These insights are critical for designing targeted interventions and enhancing safety strategies in high-risk agricultural environments.

## 4. Conclusion

This study demonstrated the potential of machine learning models, particularly ensemble methods, in predicting the severity of agricultural injuries and uncovering critical contributing factors using explainable artificial intelligence. By utilizing a dataset from AgInjuryNews (2015–2024), covering 2,421 incidents for all over the US, and examining various ML algorithms, we provided a comprehensive evaluation of predictive performance and feature importance. Ensemble models, including Random Forest, XGBoost, and LightGBM, consistently outperformed ensemble ML models in terms of accuracy and recall, with XGBoost achieving a perfect recall for fatal injuries.

While ensemble models provided the highest accuracy, their computational cost was higher compared to simpler models like Naïve Bayes, KNN and Decision Trees. This trade-off between model complexity and performance suggests that the choice of model should be aligned with the specific needs of the application, whether real-time prediction or post-incident analysis. However, challenges in classifying non-fatal injuries due to class imbalance were observed across all models, indicating the need for further optimization in data preprocessing and model training strategies.

SHAP analysis revealed that age, gender, location, and time were the most influential features across models, offering interpretable insights into the factors driving agricultural injury severity predictions. The findings align with previous studies highlighting the role of demographic and environmental variables in injury outcomes. This transparency not only enhances model interpretability but also aids stakeholders in prioritizing interventions, such as targeted safety training for older workers, male operators, or specific locations with high injury rates.

Despite these advances, the study underscores several limitations, including the manual effort required for data extraction, the need for a more balanced dataset, and the black-box nature of certain ensemble models. Future work should focus on developing automated data extraction systems, incorporating cost-sensitive learning techniques to mitigate class imbalance, and exploring domain-specific feature engineering to improve model performance further.

The practical implications of this research are significant. By combining high-performance predictive models with explainable AI, stakeholders in agricultural safety can better understand injury risk factors, design effective preventive strategies, and allocate resources more efficiently. This study serves as a foundation for integrating advanced ML techniques into agricultural safety practices, ultimately contributing to reducing injury rates and enhancing the well-being of agricultural workers.

## Data Availability

All data produced in the present study are available upon reasonable request to the authors

